# Single-cell mtDNA heteroplasmy in colorectal cancer

**DOI:** 10.1101/2021.11.24.21266805

**Authors:** João Almeida, Andrés Pérez-Figueroa, João M. Alves, Monica Valecha, Sonia Prado-López, Pilar Alvariño, Jose Manuel Cameselle-Teijeiro, Debora Chantada, Miguel M. Fonseca, David Posada

## Abstract

Human mitochondria can be genetically distinct within the same individual, a phenomenon known as heteroplasmy. In cancer, this phenomenon seems exacerbated, and most mitochondrial mutations seem to be heteroplasmic. How this genetic variation is arranged within and among normal and tumor cells is not well understood. To address this question, here we sequenced single-cell mitochondrial genomes from multiple normal and tumoral locations in four colorectal cancer patients. Our results suggest that single cells, both normal and tumoral, can carry various mitochondrial haplotypes. Remarkably, this intra-cell heteroplasmy can arise before tumor development and be maintained afterward in specific tumoral cell subpopulations. At least in the colorectal patients studied here, the somatic mutations in the single-cells do not seem to have a prominent role in tumorigenesis.

## Introduction

As each human cell contains hundreds or thousands of mitochondria (Stewart and Chinnery 2021), wild-type and mutant mtDNA can co-exist in a state called *heteroplasmy*. Heteroplasmy levels can change within a given cell due to different processes like relaxed replication, degradation, *de novo* mutation, intercellular transfer, and recombination (Stewart and Chinnery 2015; Johnston and Burgstaller 2019). As mitochondria are randomly distributed to daughter cells during cell division, the levels of heteroplasmy among cells can also fluctuate over time (Elson et al. 2001) and modulate the potential phenotypic penetrance of associated diseases, including cancer (Wallace and Chalkia 2013; McMahon and LaFramboise 2014; Stewart and Chinnery 2015; Stefano and Kream 2016; Hopkins et al. 2017; Stefano et al. 2017; Fendt et al. 2020). Potentially, the proportion of mutant mtDNA in a tissue may drift toward fixation and reach homoplasmy. However, in cancer, the vast majority of the tumoral mtDNA mutations (>85%) in tissue samples seem to be heteroplasmic, with variant allele frequencies (VAFs) lower than 0.6 (Yuan et al. 2020). How the overall mtDNA heteroplasmy is structured among and within cells is unknown. A given amount of mtDNA heteroplasmy can be explained by a population of cells carrying each a single but distinct mtDNA haplotype (*intercellular homoplasmy*) or by a population of more or less similar cells carrying several mtDNA haplotypes each (*intracellular heteroplasmy*). In other words, we do not know whether single cells typically have one or multiple mtDNA haplotypes (He et al. 2010). Due to random drift and selection, within-cell homoplasmy is expected to take place in dividing cells eventually, but not necessarily in non-dividing cells (Pérez-Amado et al. 2021).

On the other hand, the number of mtDNA molecules per cell fluctuates among human tissues (D’Erchia et al. 2015). In many types of cancer, tumor cells have fewer copies of mtDNA than the normal cells (Tseng et al. 2006; McMahon and LaFramboise 2014; Reznik et al. 2016). Variation in the number of mtDNA copies can serve as a potential biomarker for cancer, where a higher risk was associated with copy number variation (Lan et al. 2008; Thyagarajan et al. 2013). However, how mtDNA copy number varies within and among cells has not yet been studied.

Finally, intratumor heterogeneity (ITH), particularly the spatial separation of somatic mutations within a tumor, is a determinant tumor characteristic in malignant growth, invasion, metastasis, and resistance acquisition (Bedard et al. 2013; Stanta and Bonin 2018). Nonetheless, the levels of mtDNA ITH across different samples of the same tissue have not been studied yet at the single-cell level.

This study leveraged tumor multiregional single-cell whole-genome data from four colorectal cancer patients to overview how mtDNA mutation frequency and copy number are distributed among and within cells and across geographical space.

## Results

### Single-cell mtDNA coverage heterogeneity

The average sequencing depth, or coverage, across the mitochondrial genome, was 998×, 574×, 236× and 881× for CRC01, CRC07, CRC08, and CRC12, respectively. The coverage distribution was heterogeneous, with a consistent overrepresentation of particular regions across patients (Table S1, Figure S1). The breadth of the coverage, i.e., the percentage of positions covered by at least one read, was 92-97%. Around 70% of the genomes were covered on average by at least 10 reads (only 57% for CRC08).

### Single-cell mtDNA variants

We identified 3, 16, 23, and 20 mtDNA variants, primarily single nucleotide changes, in patients CRC01, CRC07, CRC08, and CRC12, respectively (Table S2). In CRC01, we found 3 germline variants. In CRC07, we identified 2 germline, 4 somatic, and 10 somatic/germline variants. In CRC08, we found 2 germline, 7 somatic, and 14 somatic/germline variants. Finally, in CRC12, we identified 13 germline and 7 somatic/germline variants. See Table S3 for a definition of the somatic, germline, and somatic/germline categories.

### Within-cell mtDNA heteroplasmy

We found no heteroplasmic variants in CRC01 (Figure 1), but at least one in 60/105 cells (57.1%) in CRC07 (Figure 2), 35/52 (67.3%) in CRC08 (Figure 3), and 9/65 (13.8%) in CRC12 (Figure 4). In CRC07, we observed within-cell heteroplasmy (VAF between 0.1 and 0.9) in 13/16 (81.3%) variant sites, and among-cell heteroplasmy (i.e., VAF less than 0.1 in some cells and more than 0.9 in others) in 8/16 (50%) (Figure 2). At site 15149, a somatic tumor variant appears at high frequency in the tumoral bulk and in specific tumor cells sampled from the distal and middle regions while is absent from the healthy cells. At site 6220, multiple normal and tumor cells showed a somatic variant at a shallow frequency –except for a normal cell, which shows a very high VAF– that was not detected in the corresponding bulk samples. At site 13993, we see a similar pattern, with a few normal and tumor cells showing a fixed somatic variant. At site 10704, a high-frequency variant was absent in some cells and fixed in others regardless of their normal/tumor status. At site 13966, we observed a similar situation, although in this case, the variant was fixed in both normal and tumoral bulk samples.

**Figure 1.**
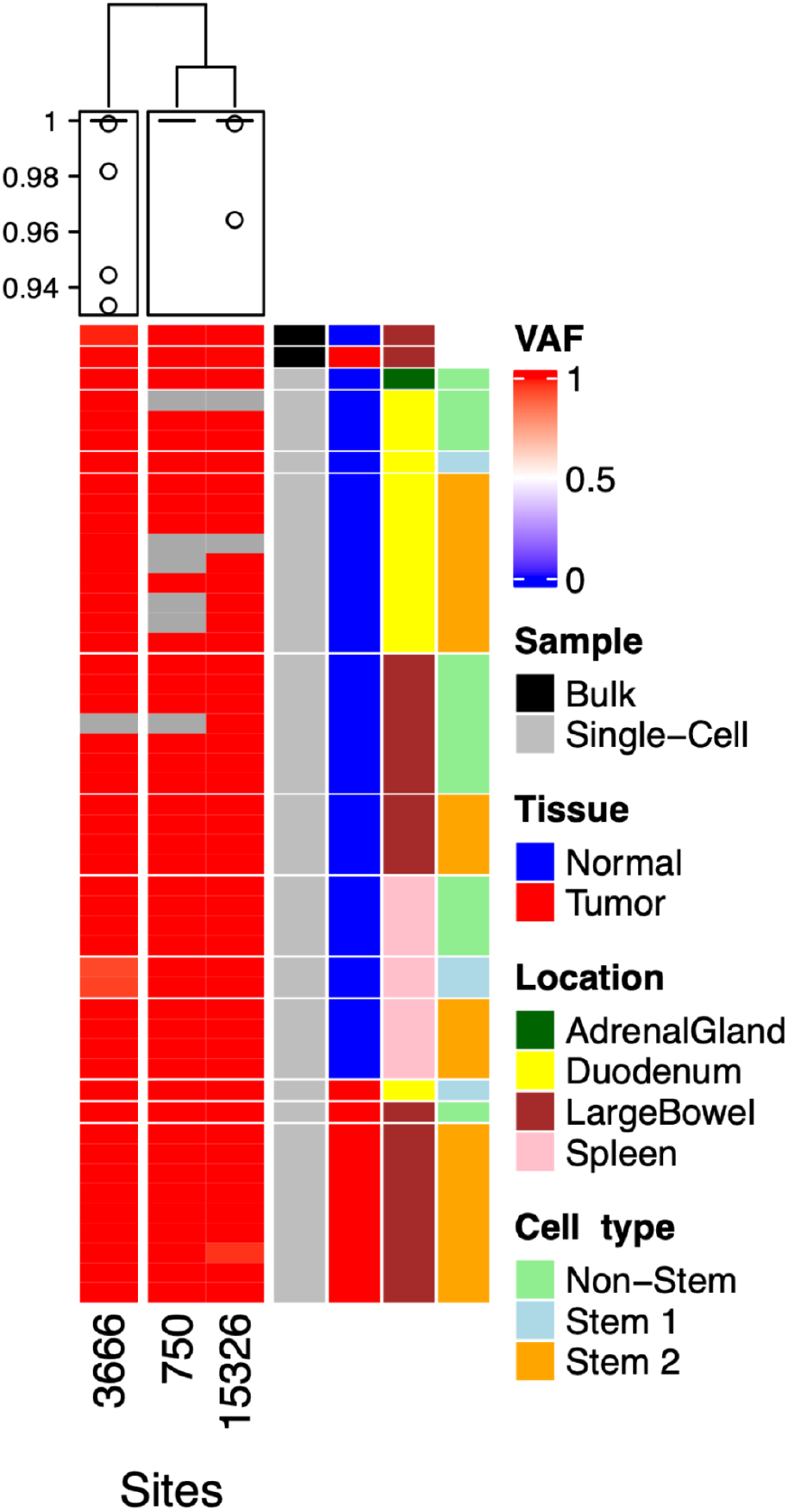
Single-cell mtDNA variant allele frequencies for CRC01. The plot depicts the mtDNA variant allele frequencies (VAFs) for two bulk samples (on top, one normal and one tumoral) and 46 single-cells at three variable sites in patient CRC01. VAF values vary from blue to red, representing the extremes 0 and 1, and grey indicates missing data (no reads at that site). The four rightmost columns indicate which rows represent bulk or single-cell samples, normal or tumoral tissue, different anatomical locations, and distinct cell types.

**Figure 2.**
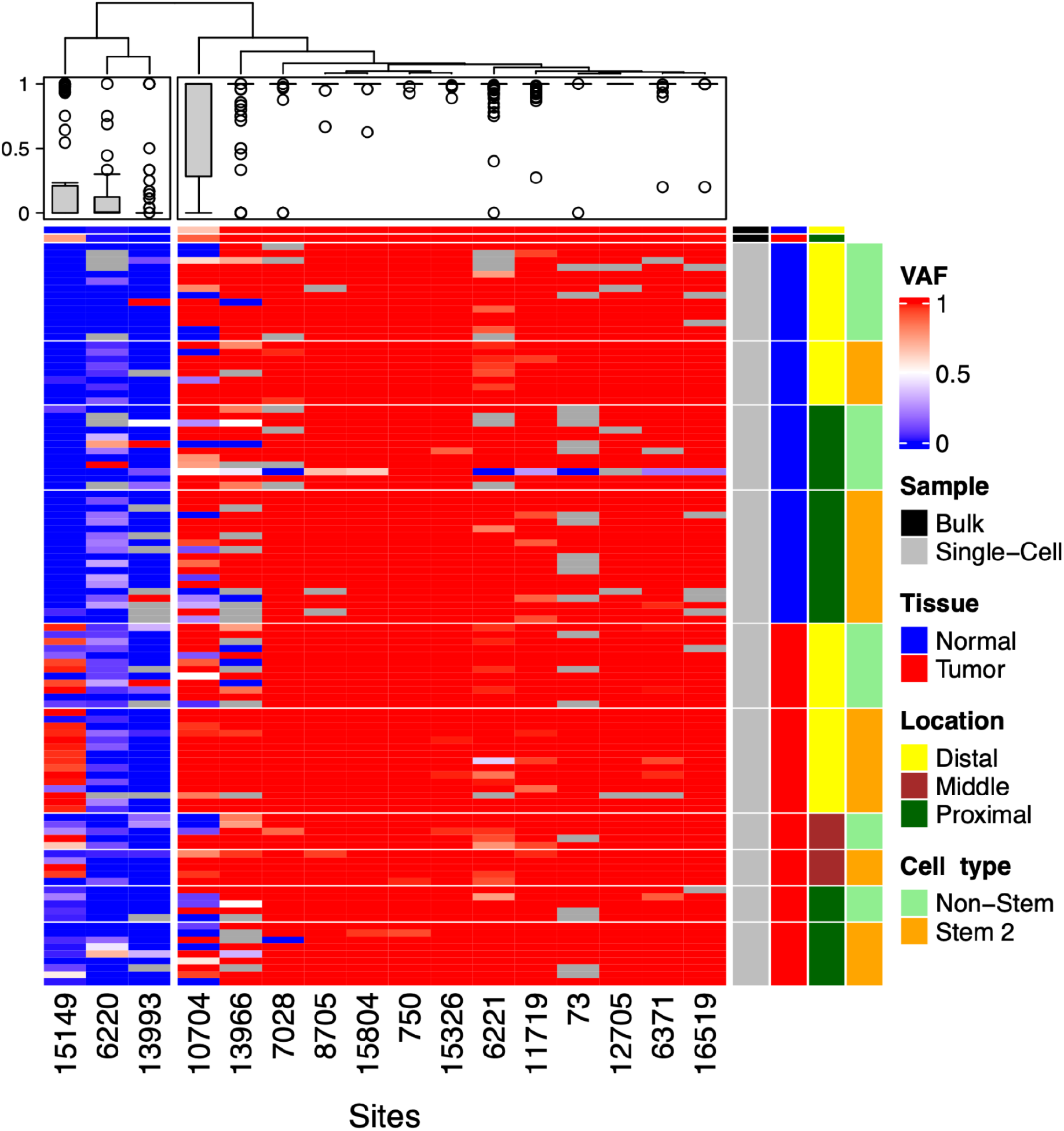
Single-cell mtDNA variant allele frequencies for CRC07. The plot depicts the mtDNA variant allele frequencies (VAFs) for two bulk samples (on top, one normal and one tumoral) and 105 single-cells at 16 variable sites in patient CRC07. VAF values vary from blue to red, representing the extremes 0 and 1, and grey indicates missing data (no reads at that site). The four rightmost columns indicate which rows represent bulk or single-cell samples, normal or tumoral tissue, different anatomical locations, and distinct cell types.

**Figure 3.**
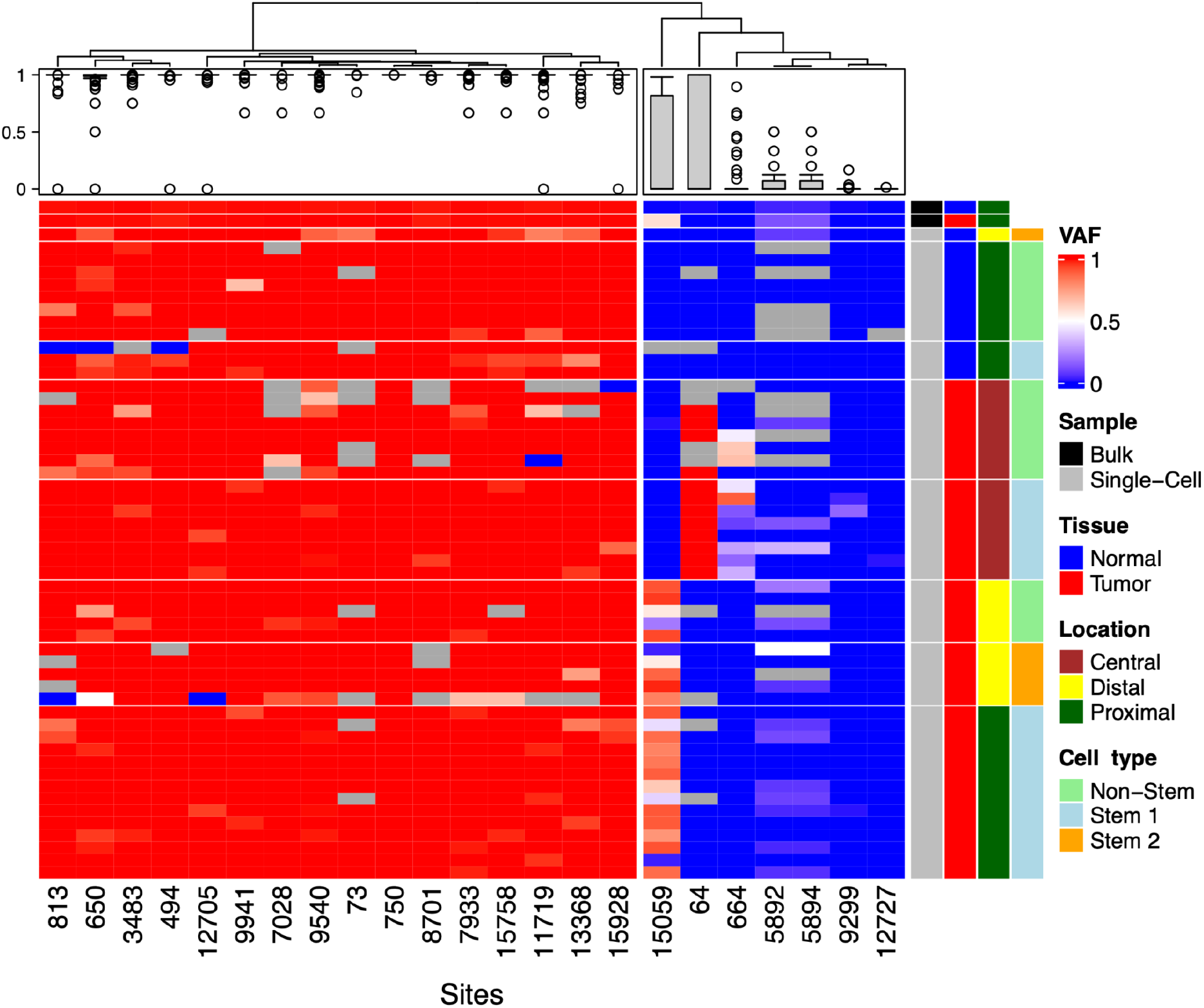
Single-cell mtDNA variant allele frequencies for CRC08. The plot depicts the mtDNA variant allele frequencies (VAFs) for two bulk samples (on top, one normal and one tumoral) and 52 single cells at 23 variable sites in patient CRC08. VAF values vary from blue to red, representing the extremes 0 and 1, and grey indicates missing data (no reads at that site). The four rightmost columns indicate which rows represent bulk or single-cell samples, normal or tumoral tissue, different anatomical locations, and distinct cell types.

**Figure 4.**
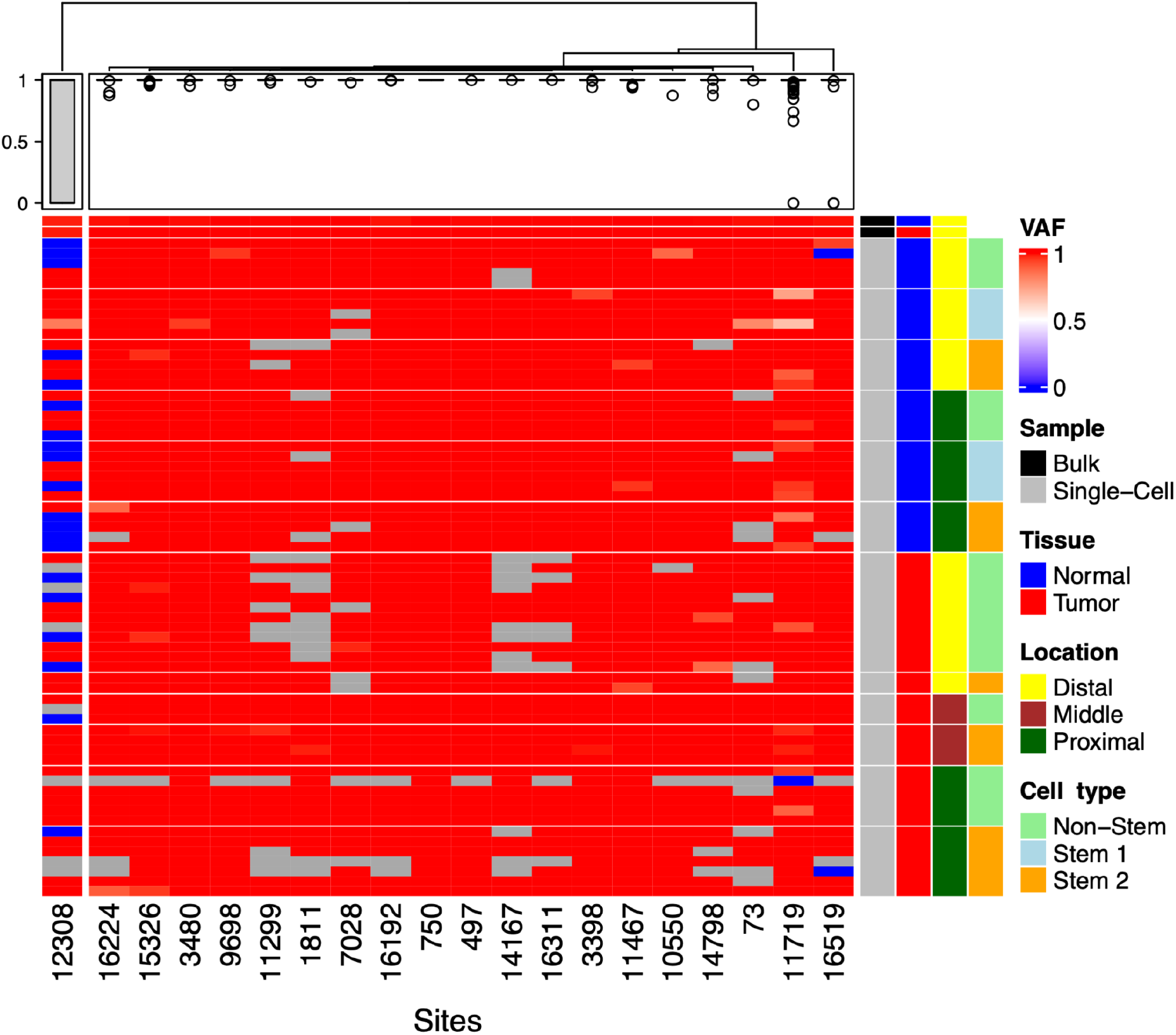
Single-cell mtDNA variant allele frequencies for CRC12. The plot depicts the mtDNA variant allele frequencies (VAFs) for two bulk samples (on top, one normal and one tumoral) and 65 single cells at 20 variable sites in patient CRC12. VAF values vary from blue to red, representing the extremes 0 and 1, and grey indicates missing data (no reads at that site). The four rightmost columns indicate which rows represent bulk or single-cell samples, normal or tumoral tissue, different anatomical locations, and distinct cell types.

In CRC08, we observed within-cell heteroplasmy in 17/23 (73.9%) variants and among-cell heteroplasmy in 8/23 (34.8%) (Figure 3). At sites 64 and 664, the alternative allele only appears in tumoral cells sampled from the central region, reaching complete fixation at site 64. At site 15059, all normal cells and the tumor cells sampled from the central area are homoplasmic for the reference allele, while most tumor cells from the proximal and distal regions show a high VAF. In CRC12, we observed within-cell heteroplasmy in 6/20 (30%) variants and among-cell heteroplasmy in 3/20 (15%) (Figure 4). The variant at site 12308 was absent in some cells and fixed in others regardless of their normal/tumor status.

### Functional impact of single-cell mtDNA variants

Heteroplasmic sites were significantly enriched in missense mutations in CRC07, stop gains in CRC08, and rRNA mutations in CRC12 (Table S4). The number of non-synonymous and synonymous mutations in normal and tumor cells was too small for a reliable estimation of the ratio of non-synonymous to synonymous rates (dN/dS) (Nei and Gojobori 1986). Non-synonymous variants did not have, overall, a higher VAF than synonymous variants in the tumors.

### Single-cell mtDNA population structure

The FST statistic for cell differentiation was relatively high for CRC07, CRC08, and CRC12 (0.729, 0.667, and 0.878, respectively), but not for CRC01 (0.079). These FST values suggest that a larger fraction of the observed heteroplasmy results from differences among cells. Still, at the same time, there is a noticeable level of within-cell heteroplasmy, as we can appreciate in the VAF plots. The analysis of molecular variance (AMOVA) unveiled significant genetic differences among normal and tumor cell populations, among anatomical locations, and among cell types in CRC07 and among anatomical areas in CRC08 (Table 1). We could not run the AMOVA for patient CRC01 because of its low genetic variability.

**Table 1.** Analysis of Molecular Variance (AMOVA). P-values of the AMOVA for the Nei’s genetic distances computed from the single-cell VAF values across all individual samples and for different hierarchical levels.

| Group | CRC07 | CRC08 | CRC12 |
| --- | --- | --- | --- |
| Tumor/Normal | **0.000\*\*\*** | 0.394 | 0.971 |
| Location within Tumor/Normal | **0.001\*\*\*** | **0.000\*\*\*** | 0.149 |
| Cell type within Tumor/Normal | **0.019\*** | 0.055 | 0.535 |

\* p-value $\leq 0.05$ , \*\* p-value $\leq 0.01$ , \*\*\* p-value $\leq 0.001$

In addition, we found significant pairwise genetic differences in CRC07 between all tumoral locations (Table S5) and between two of the tumoral areas and the normal locations. We also found significant differences in CRC08 between the normal proximal location and all three tumoral locations and between the central tumor location and the other two tumor locations. In CRC12, only the distance between the distal and proximal tumor locations was significant. Besides, we observed significant pairwise genetic differences in CRC07 among every cell type, except among tumor stem 2 (TS2) and tumor non-stem cells (TNS) (Table S6). In CRC08, we found significant differences between several non-stem and stem cell types in normal and tumor tissues. Finally, in CR12, we only observed significant differences between TS2 and normal stem 2 (NS2) cells.

We carried out principal component analyses to better characterize the mtDNA differences among the different groups of cells. The separation between specific tumoral and normal cells was not apparent in any of the patients (Figure S2), although in CRC08, the tumor cells are much more dispersed than the normal cells. When considering the geographical location, most cells overlap in the four patients (Figure S3). The same lack of specific patterns can be seen at the cell type level (Figure S4).

### Single-cell mtDNA copy number variation

We estimated a statistically significant (p-value ≤ 0.05) higher number of mtDNA copies in tumor cells in patients CRC01 (16507 copies on average in tumor cells versus 2197 in normal cells), CRC07 (302 vs. 181), and CRC12 (534 vs. 258), but not in CRC08 (135 vs 167; p-value = 0.48) (Figure 5). In CRC01, we observed a significantly higher number of mtDNA copies in the large bowel’s normal cells than in the duodenum. In CRC07, we detected significantly more mtDNA copies in normal and tumor cells in the distal region than in the proximal region.

**Figure 5.**
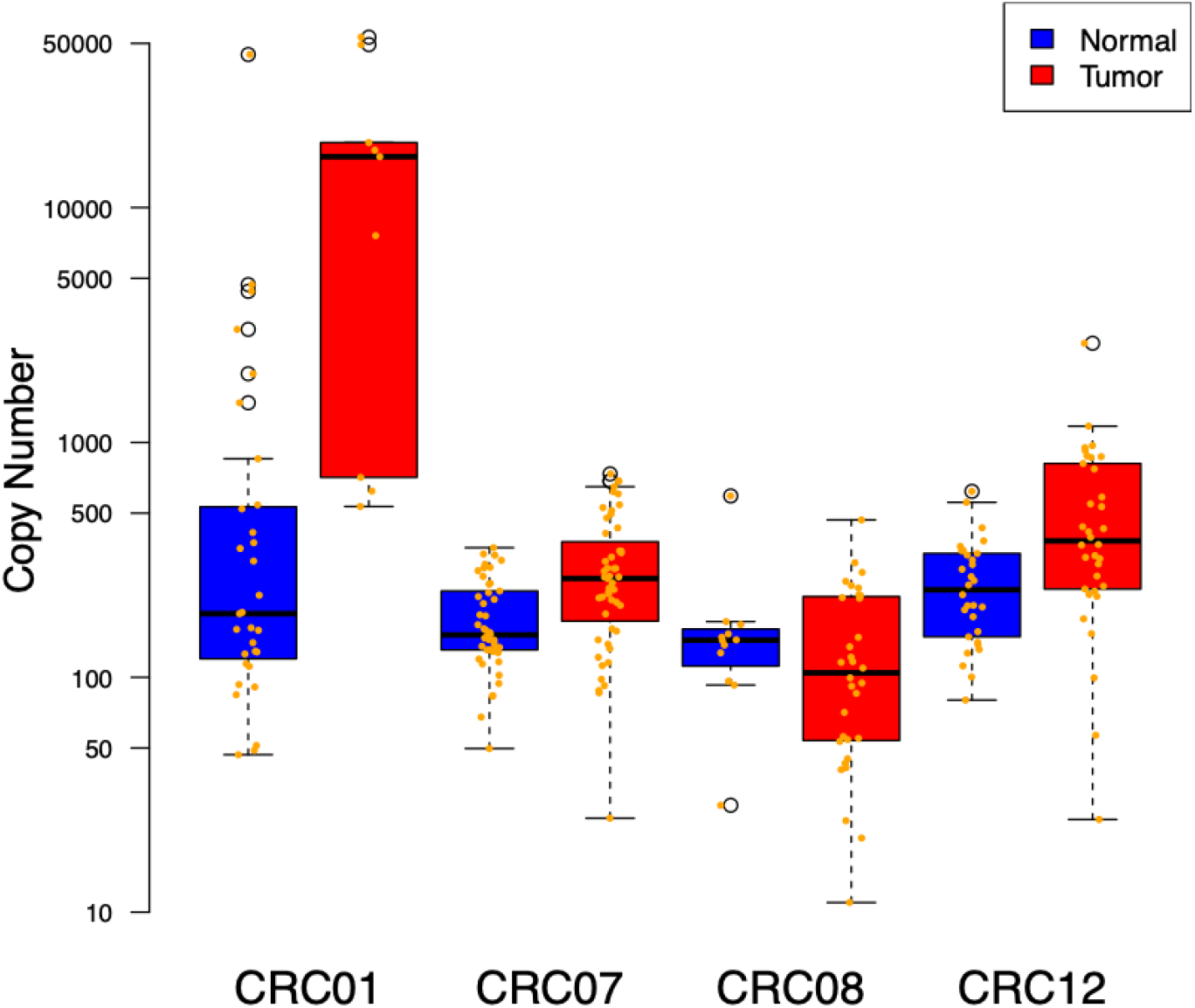
Single-cell mtDNA copy number in normal and tumor cells. Copy number for some cells could not be calculated due to a lack of diploid regions (CRC01, n= 40; CRC07, n= 96; CRC08, n= 44; CRC012, n= 64).

## Discussion

In this study, we have leveraged single-cell whole-genome sequencing data to study the levels of mtDNA heteroplasmy within and between cells in four CRC patients. To our knowledge, this is the first study addressing mtDNA heteroplasmy at the single-cell level in normal and cancer tissues.

We identified a limited number of somatic mtDNA variants, consistent with previous studies (Yuan et al. 2020). Still, our results indicate that a single (normal or tumoral) cell can carry multiple mtDNA haplotypes and that these levels of intra-cell heteroplasmy can change from patient to patient.

Notably, at least in patients CRC07 and CRC08, some sites are heteroplasmic both in normal and tumor cells, suggesting that the genetic bottleneck produced during transformation (i.e., we assume that all the tumor cells descend from a single, ancestral tumor cell) does not necessarily eliminate the intra-cell mtDNA variation present in the transformed cell. In other words, this suggests not only that many mtDNA mutations do pre-date cancer itself (Chinnery et al. 2002) but also that these mutations can be heteroplasmic within the original tumor cell.

In our data set, heteroplasmy is mainly a result of differences among cells than within cells, as expected in a population of dividing cells where the mitochondrial population is subject to drift and, potentially, selection. Still, intracell heteroplasmy does occur. We observed some changes in within-cell heteroplasmy among normal and tumor cell populations, among distinct anatomical locations, or different cell types in some patients. In the VAF plots, we could identify clear groups of cells with consistent allele frequency changes at specific sites. For example, we observed three sites in which a variant was absent in the normal cells but frequent in at least some tumor regions (T15149C in CRC07 and at C64T and G15059A in CRC08). Curiously, in CRC08, these two variants reached fixation (or almost fixation) in different regions of the same tumor. Still, these VAF changes do not appear to have a clear functional relevance. Skonieczna et al. (2018) found several mtDNA variants in a cohort of 100 CRC patients that were heteroplasmic in the normal tissue but homoplasmic in the tumors. Still, we did not observe cases like this, perhaps because of our limited sample size.

We estimated a higher mtDNA copy number in the tumoral than in normal cells. Despite mitochondrial copy numbers varying wildly within and across cancers (Yuan et al. 2020), our numbers are consistent with the increased mtDNA copy number seen in several types of cancer (Tickoo et al. 2000; Lan et al. 2008; Shen et al. 2010; Thyagarajan et al. 2013).

In principle, mtDNA somatic mutations accumulate in a neutral, or nearly neutral, fashion (Ju et al. 2014; Yuan et al. 2020). In our single-cell datasets, non-synonymous variants did not reach a higher VAF than synonymous variants, suggesting in any case that the variants we detected are not associated with tumor development.

## Methods

### Sample collection

We obtained multiple tumoral and normal tissue samples from four colorectal cancer patients (Table S7). CRC01 samples were obtained during a warm autopsy, while the samples from the other patients were obtained from excess tumor tissue present in the colectomy specimens. All colorectal cancers were conventional adenocarcinomas (not otherwise specified), according to the criteria of the latest World Health Organization (WHO) digestive system tumors classification (Who Classification of Tumours Editorial Board 2019). Samples included in this study were provided by the Biobanks of the Health Research Institute of Santiago (PT13/0010/0068) and Galicia Sur Health Research Institute (B.0000802), both integrated into the Spanish National Biobank Network. Samples were processed following standard operating procedures with the approval of the Ethical and Scientific Committees (CAEI Galicia 2014/015). Written informed consents were provided by the patients or by their families.

### Tumor disaggregation and sorting

We froze the tissue samples in liquid nitrogen, placed them in dry ice, and transported them to the laboratory. Next, we minced the samples into pieces of 1 mm^3^ with a scalpel and digested by incubation in Accutase (LINUS) for 1 h at 37 °C. After that, we filtered the cell suspension with a 70 μm cell strainer (FALCON) and assessed cell viability with Triptan Blue (Gibco). When the percentage of dead cells exceeded 30%, we did a Ficoll-Paque density gradient centrifugation to get rid of dead cells before sorting. We washed the cell pellets twice, suspended them in ice-cold phosphate-buffered saline (PBS), and stained them for 30 min with the following monoclonal antibodies: Anti-EpCAM (EBA1) (FITC)-conjugated, Anti-CD44 (APC)-conjugated, Anti-CD166 (PE)-conjugated, Anti-Lgr5 (VB 421)-conjugated. All antibodies were purchased from BD Biosciences. Following three successive washes in the PBS buffer, we added DRAQ5 and 7AAD dyes to select nucleated cells and exclude non-viable ones. We carried out flow cytometry analyses, and sorting of EpCAM+/CD44-/CD166-/Lgr5+ (tumoral stem 1 [TS1] and normal stem 1 [NS1] cells); EpCAM+/CD44+/CD166+/Lgr5- (tumoral stem 2 [TS2] and normal stem 2 [NS2]) and EpCAM+/CD44-/CD166-/Lgr5- (tumoral non-stem [TNS] and normal non-stem [NNS]) cell populations with a FACS ARIA III (BD Biosciences), and analyzed the data with the BD FACSDiva and Miltenyi Biotec Flowlogic software. In total, we selected 268 cells for further analysis (46 for CRC01, 105 for CRC07, 52 for CRC08 and 65 for CRC12), from those, 133 (18 TS1 + 59 TNS + 56 TS2) were normal (healthy) and 135 (22 TS1 + 66 TNS + 47 TS) were tumor cells (Table S7).

### Whole-genome single-cell and bulk sequencing

#### Single-cell whole genome amplification

To obtain enough DNA for sequencing from the individual cells, we carried out single-cell whole genome amplification (scWGA) with the Ampli1 Kit from Silicon Biosystems. To minimize potential contamination, we worked under a Biological Safety Cabinet, UV-irradiated all the plastic materials employed, and used a dedicated set of pipettes. In addition to patient cells, we included a positive (10 ng/µl REPLIg human control kit, QIAGEN) and negative (DNase/RNase free water) control in the amplification process. Next, we assessed the quality of the amplified DNA with the Ampli1 QC Kit and selected the positive samples for the 4 PCR DNA fragments. Then, we used the Ampli1 ReAmp/ds kit on the selected samples to increase the total double-stranded DNA. Later, we removed the kit adaptors adding 5 µl of NE Buffer 4 10X (New England Biolabs), 1 µl of MseI 50U/µl (New England Biolabs), and 19 µl of nuclease-free water to every 25 µl of a sample. We introduced the resulting mix in a thermocycler. Next, we applied a program consisting of a step of 37°C for 3 h, followed by 20 minutes at 65°C for enzyme inactivation. Then, we purified the Ampli1 products using 1.8X AMPure XP beads (Agencourt, Beckman Coulter). Later, we measured DNA yield, integrity, and amplicon size distribution with a Qubit 3.0 fluorometer (ThermoFisher Scientific) and a 2200 TapeStation platform with the D5000 ScreenTape assay (Agilent Technologies).

#### Bulk genomic DNA (gDNA) isolation

We isolated the gDNA from the bulk samples using the QIAamp DNA Mini kit (QIAGEN) following the fabricant recommendations. Next, We estimated the yield and DNA integrity as described for the single cells above but using the Genomic DNA Screentape Assay instead.

#### Library construction and next-generation sequencing

We sent all samples to the Spanish National Center for Genomic Analysis (CNAG), where bulk and single-cell whole-genome sequencing libraries were built using the KAPA (Kapa Biosystems) library preparation kit with some modifications. Bulk and single-cell libraries were sequenced at ∼39X (bulk) and ∼6X (single cells) in an Illumina NovaSeq 6000 platform.

#### Mitochondrial variant calling

To avoid the detection of mtDNA mutations caused by pseudogenes or homologous sequences in nuclear DNA (NUMTs), we aligned the reads to both nuclear and mitochondrial reference genomes, treating NUMTs and mitochondrial specific mutations equally, and thus, artificially producing a high coverage on these nuclear positions while creating a deficit in the mtDNA ones. This method, which trades less interference of NUMTs in mtDNA for lack of detection power for those positions in the mitogenome, has already been used in mtDNA variant calling pipelines (Guo et al. 2013). We mapped the reads to the Revised Cambridge Reference Sequence (rCRS) with SAMtools (Li et al. 2009) to get the first set of reads ready for calling input.

We converted back mtDNA reads mapped to rCRS and unmapped reads into FASTQ, following existing pipelines (Ding et al. 2015), and remapped them to a “shifted” rCRS, for consideration on the circularity of the mitochondrial genome (mt-genome). This procedure avoids unaligned and discarded reads on the extremities of the reference resulting from the artificial breakpoint of the mt-genome on the replication control region, defining the “start” at position 1 and “end” at position 16,569. In this double-alignment method (Ding et al. 2015), the “shifted” reference is created by switching the positions of, roughly, the first and second half of the rCRS so that positions 8001–16569 appear first, followed by positions 1–8000.

We applied quality control filters to avoid calling errors by selecting reads with a base quality score ≥ 20, median depth ≥ 100 per individual, raw depth ≥ 40, and depth after base quality score filter ≥ 10, MAF ≥ 4%, as in Ding et al. (2015). For variant calling, we used Mutect2 on multi-sample mode (Cibulskis et al. 2013) and with option --max-mnp-distance 0. The resulting variants were filtered with FilterMutectCalls. We performed the variant calling using all the bulk and single-cell samples together. We used the reads mapped to rCRS to call the variants for coordinates 4,000–12,000 and reads mapped to the “shifted” rCRS to call for coordinates 0–4,000 and 12,000–16,000. Then, we merged the two sets of called variants. Finally, we removed multiallelic variants, read depth 1, and variants with more than 20% missing data.

#### Estimation of the mtDNA copy number

We used fastMitoCalc (Ding et al. 2015; Qian et al. 2017) to estimate the mtDNA copy number as the depth ratio between mtDNA and nuclear DNA:

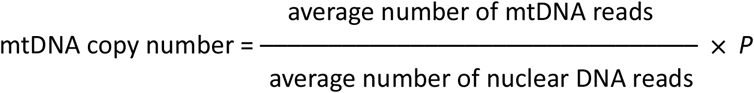

where *P* is the ploidy. This procedure assumes that regions of the genome of equal ploidy have the same depth (Guo et al. 2013; Samuels et al. 2013; Zhao et al. 2013; Cai et al. 2015; D’Erchia et al. 2015; Ding et al. 2015; Reznik et al. 2016; Zhang et al. 2017). For normal diploid cells, *P* is 2; however, cancer cells can exhibit large-scale genomic amplifications and deletions, altering their ploidy (Reznik et al. 2016). Therefore, we calculated the mtDNA copy number using only diploid nuclear regions (Cai et al. 2015; Zhang et al. 2017). We called single-cell copy-number variants (CNVs) with Ginkgo (Garvin et al. 2015) to identify these regions using variable-length bins of around 500 kb. After binning, data for each cell was normalized and segmented using default parameters.

#### Identification of within-cell heteroplasmy

We calculated the variant allele frequency (VAF) by dividing the allelic depth of the alternative allele by the total read depth. We classified as somatic those variants present in at least one single (normal or tumor) cell and absent or not fixed in the normal bulk, as germline those variants fixed in the normal bulk. Variants fixed in the normal bulk and present in at least one single (normal or tumoral) cell but not fixed were classified as germline and somatic. We consider a variant to be heteroplasmic for a given cell if its VAF was between 0.1 and 0.9. Otherwise we classified it as homoplasmic (0.1 > VAF > 0.9).

#### Population genetic structure

To quantify the structuration of heteroplasmy within and among cells, we computed the haploid equivalent of the FST statistic (Wright 1951), whose values go from 0 to 1. An FST of 0 will indicate that all heteroplasmy occurs because of differences among cells. A value of 1 will suggest that all heteroplasmy occurs because of differences within cells.

In addition, we used Nei’s (Nei 1972) distances from the VAFs with an in-house R script (https://github.com/anpefi/sc-mtDNA) to describe the mitochondrial population structure at the single-cell level. We carried out an analysis of molecular variance (AMOVA) (Excoffier et al. 1992) using the R package *pegas* (Paradis 2010) across different hierarchical levels: tumor vs. normal, location within tumor/normal, and cell type within tumor/normal. Additionally, a pairwise AMOVA, using the same method as the hierarchical AMOVA, was applied to the different groups at the location per tissue and cell type per tissue level. To evaluate the null hypothesis of no population structure, we used permutation tests with 10,000 replicates.

#### Functional analyses

To look for differences between the functional characterization of tumor and normal cells, we looked for alternatives to “typically normal allele” homoplasmic variants. We identified these “typically normal allele” homoplasmic variants as the variants that differ from those considered as individually fixed in normal cells and distributed them into functional groups: synonymous, missense, or stop gain, for protein-coding, and intergenic or rRNA for non-protein-coding. We annotated and prioritized the variants using Ensemble’s Variant Effect Predictor (VEP) (McLaren et al. 2016). In addition, we annotated each variant using the online SNV query of MITOMASTER (Brandon et al. 2009).

## Data Availability

All data produced are available online at https://github.com/anpefi/sc-mtDNA

## Data availability

Data and scripts are available at https://github.com/anpefi/sc-mtDNA.

## Acknowledgments

This work was supported by project No. 32030 awarded to M.F., co-financed by COMPETE 2020, Portugal 2020 and the EU through the ERDF, and by FCT (PTDC/BIA-EVL/32030/2017) through national funds, and by the European Research Council (ERC-617457-PHYLOCANCER awarded to D.P.) and Spanish Ministry of Science, Innovation, and Universities - MCIU (PID2019-106247GB-I00 awarded to D.P.). J.M.C.-T. is supported by grant no. ISCIII-PI19/01316 from Instituto de Salud Carlos III, State Research Agency and Ministry of Science and Innovation (Spain), with the participation of the European FEDER fund. We want to thank Sara Rocha for her helpful comments on this work.

## Supplementary Material

**Table S1.**
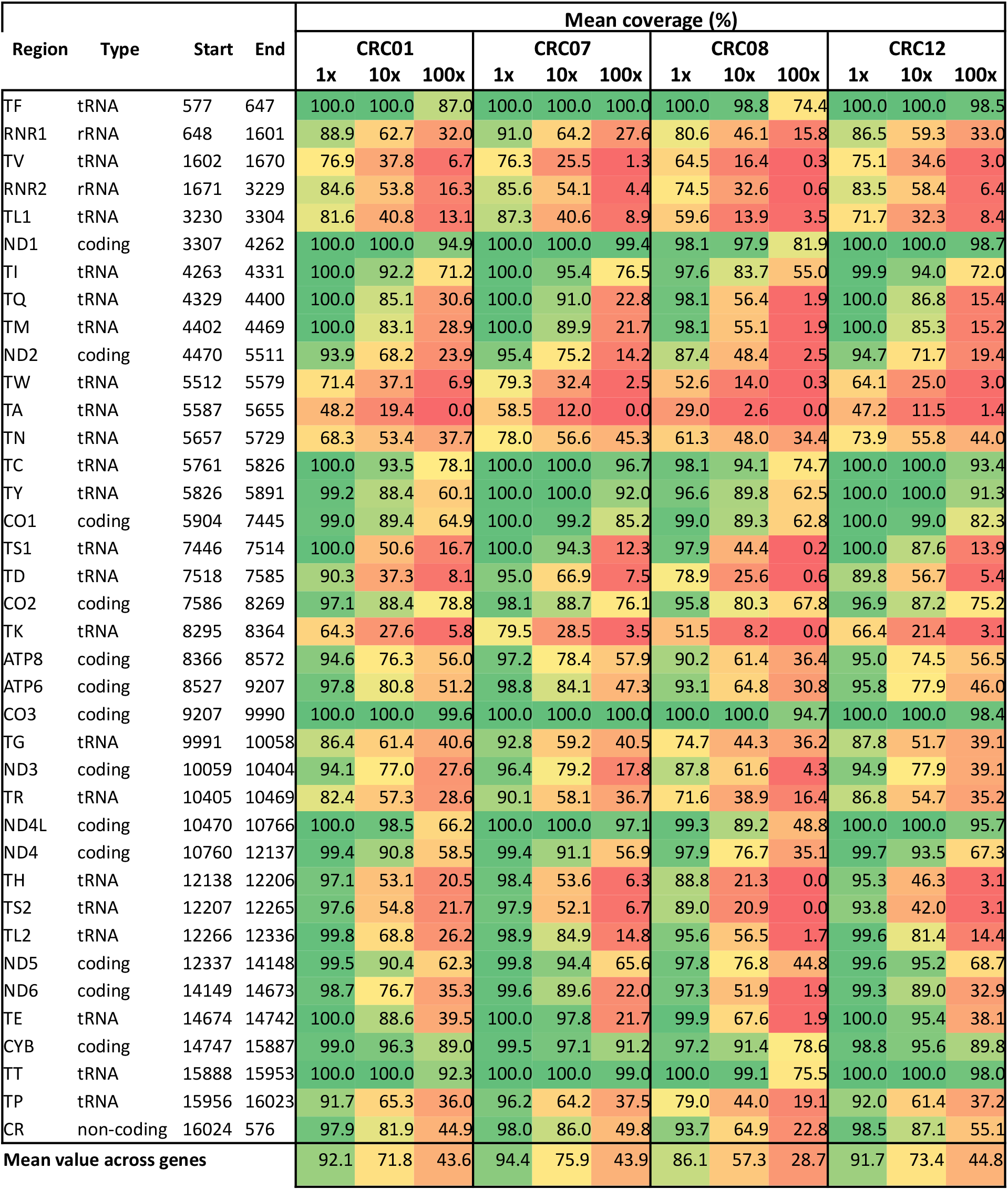
Mean individual mtDNA sequence coverage for each gene. Coverage percentage of each mitochondrial gene with at least 1, 10, and 100 reads across each individual. The bottom line presents the mean value of coverage for the overall mitogenome that expresses genes.

**Table S2.** Single-cell mtDNA variants. Var. type: variant type (see Table S3): G = Germline, S = Somatic, S/G = Somatic/Germline. Tissue Som.: tissue where the somatic variant appears N =Normal, T = Tumoral, N/T = Normal and Tumoral. Somatic variants are highlighted in bold.

| Patient | Region | Position | Change | Var. Type | Tissue Som. | Annotation | Heteroplasmy |
| --- | --- | --- | --- | --- | --- | --- | --- |
| CRC01 | RNR1 | 750 | A>G | G | -- | rRNA | No |
|  | ND1 | 3666 | G>A | G | -- | ND1:G120G | No |
|  | CYB | 15326 | A>G | G | -- | CYB:T194A | No |
| CRC07 | **CR** | **73** | **A>G** | **S/G** | **N** | **non-coding** | **Yes** |
|  | RNR1 | 750 | A>G | G | -- | rRNA | No |
|  | **CO1** | **6220** | **CT>C** | **S** | **N/T** | **frameshift** | **Yes** |
|  | **CO1** | **6221** | **T>C** | **S/G** | **N/T** | **CO1:P106P** | **Yes** |
|  | **CO1** | **6371** | **C>T** | **S/G** | **N/T** | **CO1:S156S** | **Yes** |
|  | **CO1** | **7028** | **C>T** | **S/G** | **N/T** | **CO1:A375A** | **Yes** |
|  | **ATP6** | **8705** | **T>C** | **S/G** | **N** | **ATP6:M60T** | **Yes** |
|  | **ND4L** | **10704** | **G>A** | **S** | **N/T** | **ND4L:V79I** | **Yes** |
|  | **ND4** | **11719** | **G>A** | **S/G** | **N/T** | **ND4:G320G** | **Yes** |
|  | ND5 | 12705 | C>T | G | -- | ND5:I123I | No |
|  | **ND5** | **13966** | **A>G** | **S/G** | **N/T** | **ND5:T544A** | **Yes** |
|  | **ND5** | **13993** | **C > 73bp\*** | **S** | **N/T** |  | **Yes** |
|  | **CYB** | **15149** | **T>C** | **S** | **T** | **CYB:W135R** | **Yes** |
|  | **CYB** | **15326** | **A>G** | **S/G** | **N** | **CYB:T194A** | **Yes** |
|  | **CYB** | **15804** | **T>C** | **S/G** | **N** | **CYB:V353A** | **Yes** |
|  | **CR** | **16519** | **T>C** | **S/G** | **N** | **non-coding** | **Yes** |
| CRC08 | **CR** | **64** | **C>T** | **S** | **T** | **non-coding** | **Yes** |
|  | **CR** | **73** | **A>G** | **S/G** | **N** | **non-coding** | **Yes** |
|  | **CR** | **494** | **C>T** | **S/G** | **N** | **non-coding** | **Yes** |
|  | **RNR1** | **650** | **T>C** | **S/G** | **N/T** | **rRNA** | **Yes** |
|  | **RNR1** | **664** | **G>A** | **S** | **T** | **rRNA** | **Yes** |
|  | RNR1 | 750 | A>G | G | -- | rRNA | No |
|  | **RNR1** | **813** | **A>G** | **S/G** | **N/T** | **rRNA** | **Yes** |
|  | **ND1** | **3483** | **G>A** | **S/G** | **N/T** | **ND1:E59E** | **Yes** |
|  | **NC5** | **5892** | **T>TC** | **S** | **N/T** | **non-coding** | **Yes** |
|  | **NC5** | **5894** | **A>C** | **S** | **N/T** | **non-coding** | **Yes** |
|  | **CO1** | **7028** | **C>T** | **S/G** | **T** | **CO1:A375A** | **Yes** |
|  | **CO2** | **7933** | **A>G** | **S/G** | **T** | **CO2:L116L** | **Yes** |
|  | ATP6 | 8701 | A>G | G | -- | ATP6:T59A | No |
|  | **CO3** | **9299** | **A>G** | **S** | **T** | **CO3:L31L** | **Yes** |
|  | **CO3** | **9540** | **T>C** | **S/G** | **N/T** | **CO3:L112L** | **Yes** |
|  | **CO3** | **9941** | **A>G** | **S/G** | **N/T** | **CO3:V245V** | **Yes** |
|  | **ND4** | **11719** | **G>A** | **S/G** | **N/T** | **ND4:G320G** | **Yes** |
|  | **ND5** | **12705** | **C>T** | **S/G** | **T** | **ND5:I123I** | **Yes** |
|  | ND5 | 12727 | T>C | S | T | ND5:L131L | No |
|  | **ND5** | **13368** | **G>A** | **S/G** | **N/T** | **ND5:G344G** | **Yes** |
|  | **CYB** | **15059** | **G>A** | **S** | **T** | **CYB:G105TERM** | **Yes** |
|  | **CYB** | **15758** | **A>G** | **S/G** | **N/T** | **CYB:I338V** | **Yes** |
|  | **TT** | **15928** | **G>A** | **S/G** | **T** | **tRNA** | **Yes** |
| CRC12 | **CR** | **73** | **A>G** | **S/G** | **N** | **non-coding** | **Yes** |
|  | CR | 497 | C>T | G | -- | non-coding | No |
|  | RNR1 | 750 | A>G | G | -- | rRNA | No |
|  | RNR2 | 1811 | A>G | G | -- | rRNA | No |
|  | ND1 | 3398 | T>C | G | -- | ND1:M31T | No |
|  | ND1 | 3480 | A>G | G | -- | ND1:K58K | No |
|  | CO1 | 7028 | C>T | G | -- | CO1:A375A | No |
|  | CO3 | 9698 | T>C | G | -- | CO3:L164L | No |
|  | **ND4L** | **10550** | **A>G** | **S/G** | **N** | **ND4L:M27M** | **Yes** |
|  | ND4 | 11299 | T>C | G | -- | ND4:T180T | No |
|  | ND4 | 11467 | A>G | G | -- | ND4:L236L | No |
|  | **ND4** | **11719** | **G>A** | **S/G** | **N/T** | **ND4:G320G** | **Yes** |
|  | **TL2** | **12308** | **A>G** | **S/G** | **N/T** | **tRNA** | **Yes** |
|  | ND6 | 14167 | C>T | G | -- | ND6:E169E | No |
|  | **CYB** | **14798** | **T>C** | **S/G** | **T** | **CYB:F18L** | **Yes** |
|  | CYB | 15326 | A>G | G | -- | CYB:T194A | No |
|  | CR | 16192 | C>T | G | -- | non-coding | No |
|  | **CR** | **16224** | **T>C** | **S/G** | **N/T** | **non-coding** | **Yes** |
|  | CR | 16311 | T>C | G | -- | non-coding | No |
|  | **CR** | **16519** | **T>C** | **S/G** | **N/T** | **non-coding** | **Yes** |

\* indel 73 bp: C > CCATTTGATTCCATTGATGTTGATTCCATTTCGATTCCATTATATGATGATTCCATGCAATTCCATTAGATGA

**Table S3.** Classification of variants as somatic, germline, and somatic/germline.

| **Type** | **Tissue** | **Normal bulk** | **Tumor bulk** | **Normal single-cell** | **Tumor single-cell** |
| --- | --- | --- | --- | --- | --- |
| Somatic | Tumor | absent | absent or present | absent | at least 1 cell |
| Somatic | Normal | not fixed | absent | at least 1 cell | absent |
| Somatic | Tumor and Normal | not fixed | absent or present | at least 1 cell | at least 1 cell |
| Germline | Tumor and Normal | fixed | fixed | fixed | fixed |
| Somatic / Germline | Tumor | fixed | absent or present | fixed | at least 1 cell but not fixed |
| Somatic / Germline | Normal | fixed | fixed | at least 1 cell but not fixed | fixed |
| Somatic / Germline | Tumor and Normal | fixed | absent or present | at least 1 cell but not fixed | at least 1 cell but not fixed |

**Table S4.** Functional impact of single-cell mtDNA variants.

| Patient ID | Site | Region | Consequence | Heteroplasmy |
| --- | --- | --- | --- | --- |
| CRC08 | 64 | CR | Intergenic | Yes |
| CRC07 | 73 | CR | Intergenic | Yes |
| CRC08 |  |  |  | Yes |
| CRC12 |  |  |  | Yes |
| CRC08 | 494 | CR | Intergenic | Yes |
| CRC12 | 497 | CR | Intergenic | No |
| CRC08 | 650 | RNR1 | Non-coding | Yes |
| CRC08 | 664 | RNR1 | Non-coding | Yes |
| CRC01 | 750 | RNR1 | Non-coding | No |
| CRC07 |  |  |  | No |
| CRC08 |  |  |  | No |
| CRC12 |  |  |  | No |
| CRC08 | 813 | RNR1 | Non-coding | Yes |
| CRC12 | 1811 | RNR2 | Non-coding | No |
| CRC12 | 3398 | ND1 | Missense | No |
| CRC12 | 3480 | ND1 | Synonymous | No |
| CRC08 | 3483 | ND1 | Synonymous | Yes |
| CRC01 | 3666 | ND1 | Synonymous | No |
| CRC08 | 5892 | NC5 | Non-coding | Yes |
| CRC08 | 5894 | NC5 | Intergenic | Yes |
| CRC07 | 6220 | CO1 | Frameshift | Yes |
| CRC07 | 6221 | CO1 | Synonymous | Yes |
| CRC07 | 6371 | CO1 | Synonymous | Yes |
| CRC07 | 7028 | CO1 | Synonymous | Yes |
| CRC08 |  |  |  | Yes |
| CRC12 |  |  |  | No |
| CRC08 | 7933 | CO2 | Synonymous | Yes |
| CRC08 | 8701 | ATP6 | Missense | No |
| CRC07 | 8705 | ATP6 | Missense | Yes |
| CRC08 | 9299 | CO3 | Synonymous | Yes |
| CRC08 | 9540 | CO3 | Synonymous | Yes |
| CRC12 | 9698 | CO3 | Synonymous | No |
| CRC08 | 9941 | CO3 | Synonymous | Yes |
| CRC12 | 10550 | ND4L | Synonymous | Yes |
| CRC07 | 10704 | ND4L | Missense | Yes |
| CRC12 | 11299 | ND4 | Synonymous | No |
| CRC12 | 11467 | ND4 | Synonymous | No |
| CRC07 | 11719 | ND4 | Synonymous | Yes |
| CRC08 |  |  |  | Yes |
| CRC12 |  |  |  | Yes |
| CRC12 | 12308 | TL2 | Non-coding | Yes |
| CRC07 | 12705 | ND5 | Synonymous | No |
| CRC08 | 12705 | ND5 | Synonymous | Yes |
| CRC08 | 12727 | ND5 | Synonymous | No |
| CRC08 | 13368 | ND5 | Synonymous | Yes |
| CRC07 | 13966 | ND5 | Missense | Yes |
| CRC07 | 13993 | ND5 | - | Yes |
| CRC12 | 14167 | ND6 | Synonymous | No |
| CRC12 | 14798 | CYB | Missense | Yes |
| CRC08 | 15059 | CYB | Stop gained | Yes |
| CRC07 | 15149 | CYB | Missense | Yes |
| CRC01 | 15326 | CYB | Missense | No |
| CRC07 |  |  |  | Yes |
| CRC12 |  |  |  | No |
| CRC08 | 15758 | CYB | Missense | Yes |
| CRC07 | 15804 | CYB | Missense | Yes |
| CRC08 | 15928 | TT | Non-coding | Yes |
| CRC12 | 16192 | CR | Intergenic | No |
| CRC12 | 16224 | CR | Intergenic | Yes |
| CRC12 | 16311 | CR | Intergenic | No |
| CRC07 | 16519 | CR | Intergenic | Yes |
| CRC12 |  |  |  | Yes |

**Table S5.** Pairwise mtDNA differentiation among locations. Permutation p-values of the pairwise AMOVA calculation for the Nei’s genetic distances calculated with the VAF values for different location groups per tissue. CRC01 tests were all non-significant.

|  |  |  |  |  |
| --- | --- | --- | --- | --- |
| **CRC07** | **ND** | **NP** | **TD** | **TM** |
| **NP** | 0.692 |  |  |  |
| **TD** | **0.000\*\*\*** | **0.000\*\*\*** |  |  |
| **TM** | **0.034\*** | 0.063 | **0.029\*** |  |
| **TP** | 0.956 | 0.801 | **0.000\*\*\*** | **0.043\*** |
| **CRC08** | **ND** | **NP** | **TC** | **TD** |
| **NP** | 0.328 |  |  |  |
| **TC** | 0.346 | **0.000\*\*\*** |  |  |
| **TD** | 0.277 | **0.000\*\*\*** | **0.000\*\*\*** |  |
| **TP** | 0.068 | **0.000\*\*\*** | **0.000\*\*\*** | 0.915 |
| **CRC12** | **ND** | **NP** | **TD** | **TM** |
| **NP** | 0.396 |  |  |  |
| **TD** | 0.463 | 0.258 |  |  |
| **TM** | 0.855 | 0.146 | 0.345 |  |
| **TP** | 0.915 | 0.316 | **0.010\*\*** | 0.883 |

<sup>NS</sup> non-significant; \* p-value ≤ 0.05, \*\* p-value ≤ 0.01, \*\*\* p-value ≤ 0.001

**Table S6.**
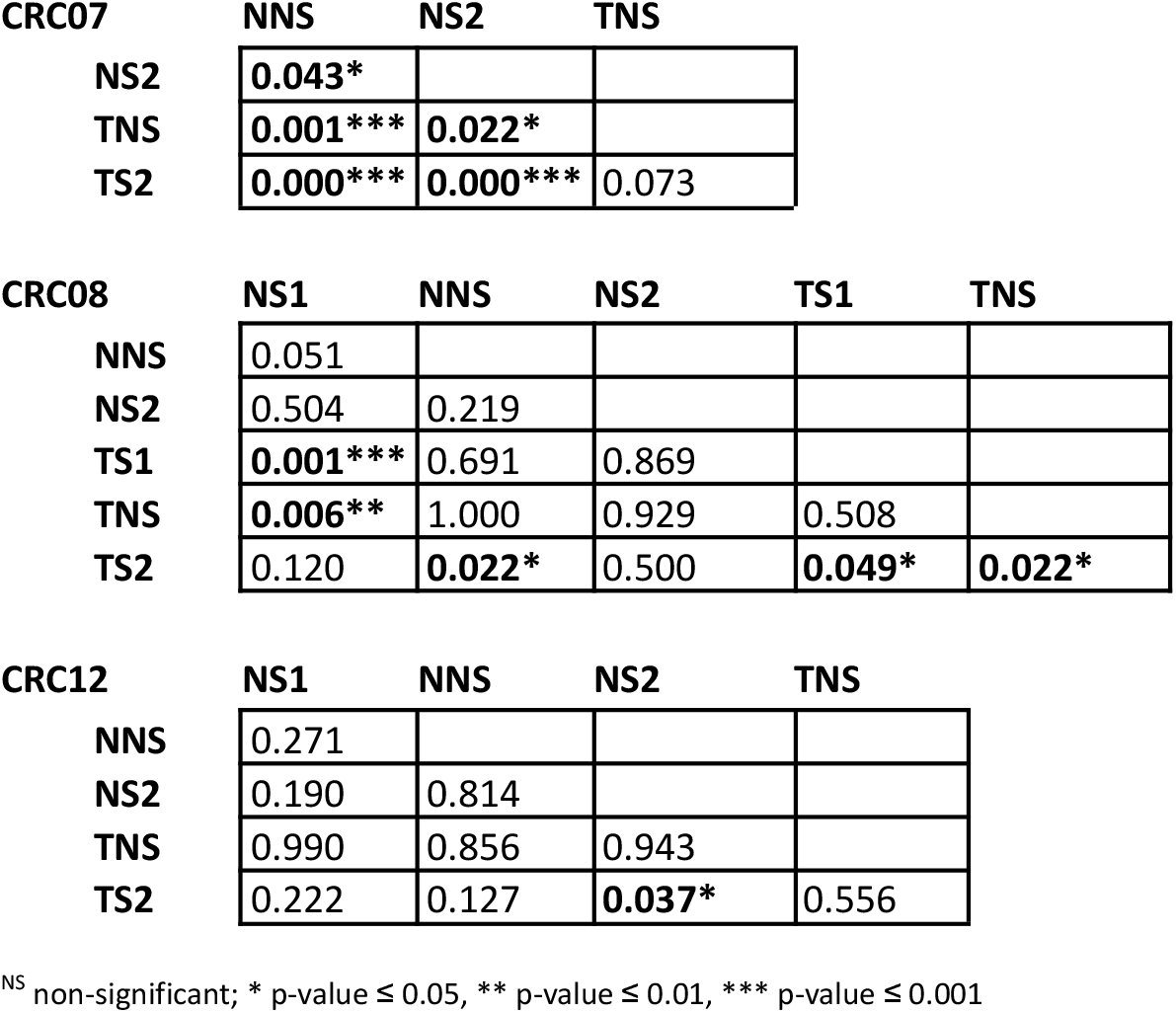
Pairwise differentiation on type per tissue. P-values of the pairwise AMOVA calculation for the Nei’s genetic distances were calculated with the VAF values for different groups of cell types per tissue.

|  |  |  |  |
| --- | --- | --- | --- |
| **CRC07** | **NNS** | **NS2** | **TNS** |
| **NS2** | **0.043\*** |  |  |
| **TNS** | **0.001\*\*\*** | **0.022\*** |  |
| **TS2** | **0.000\*\*\*** | **0.000\*\*\*** | 0.073 |

|  |  |  |  |  |  |
| --- | --- | --- | --- | --- | --- |
| **CRC08** | **NS1** | **NNS** | **NS2** | **TS1** | **TNS** |
| **NNS** | 0.051 |  |  |  |  |
| **NS2** | 0.504 | 0.219 |  |  |  |
| **TS1** | **0.001\*\*\*** | 0.691 | 0.869 |  |  |
| **TNS** | **0.006\*\*** | 1.000 | 0.929 | 0.508 |  |
| **TS2** | 0.120 | **0.022\*** | 0.500 | **0.049\*** | **0.022\*** |

|  |  |  |  |  |
| --- | --- | --- | --- | --- |
| **CRC12** | **NS1** | **NNS** | **NS2** | **TNS** |
| **NNS** | 0.271 |  |  |  |
| **NS2** | 0.190 | 0.814 |  |  |
| **TNS** | 0.990 | 0.856 | 0.943 |  |
| **TS2** | 0.222 | 0.127 | **0.037\*** | 0.556 |

<sup>NS</sup> non-significant; \* p-value $\leq 0.05$ , \*\* p-value $\leq 0.01$ , \*\*\* p-value $\leq 0.001$

**Table S7.** Single-cell samples. Cell type, geographical location, and number sampled for each patient.

| Patient | Cell Type | Geographical location | Number of cells |
| --- | --- | --- | --- |
| CRC01 | NS1 | Mucosa, duodenum | 2 |
|  | NS1 | Spleen | 2 |
|  | NNS | Adrenal gland (left) | 1 |
|  | NNS | Large bowel | 7 |
|  | NNS | Mucosa, duodenum | 3 |
|  | NNS | Spleen | 4 |
|  | NS2 | Large bowel | 4 |
|  | NS2 | Mucosa, duodenum | 9 |
|  | NS2 | Spleen | 4 |
|  | TNS | Large bowel | 10 |
| **Total** |  |  | 46 |
| CRC07 | NNS | Large bowel distal | 14 |
|  | NNS | Large bowel proximal | 12 |
|  | NS2 | Large bowel distal | 9 |
|  | NS2 | Large bowel proximal | 19 |
|  | TNS | Large bowel distal | 12 |
|  | TNS | Large bowel middle | 5 |
|  | TNS | Large bowel proximal | 5 |
|  | TS2 | Large bowel distal | 15 |
|  | TS2 | Large bowel middle | 5 |
|  | TS2 | Large bowel proximal | 9 |
| **Total** |  |  | 105 |
| CRC08 | NS1 | Large bowel proximal | 3 |
|  | NNS | Large bowel proximal | 8 |
|  | NS2 | Large bowel distal | 1 |
|  | TS1 | Large bowel central | 8 |
|  | TS1 | Large bowel proximal | 14 |
|  | TNS | Large bowel central | 8 |
|  | TNS | Large bowel distal | 5 |
|  | TS2 | Large bowel distal | 5 |
| **Total** |  |  | 52 |
| CRC12 | NS1 | Large bowel distal | 5 |
|  | NS1 | Large bowel proximal | 6 |
|  | NNS | Large bowel distal | 5 |
|  | NNS | Large bowel proximal | 5 |
|  | NS2 | Large bowel distal | 5 |
|  | NS2 | Large bowel proximal | 5 |
|  | TNS | Large bowel distal | 12 |
|  | TNS | Large bowel middle | 3 |
|  | TNS | Large bowel proximal | 6 |
|  | TS2 | Large bowel distal | 2 |
|  | TS2 | Large bowel middle | 4 |
|  | TS2 | Large bowel proximal | 7 |
| **Total** |  |  | 65 |

**Figure S1.**
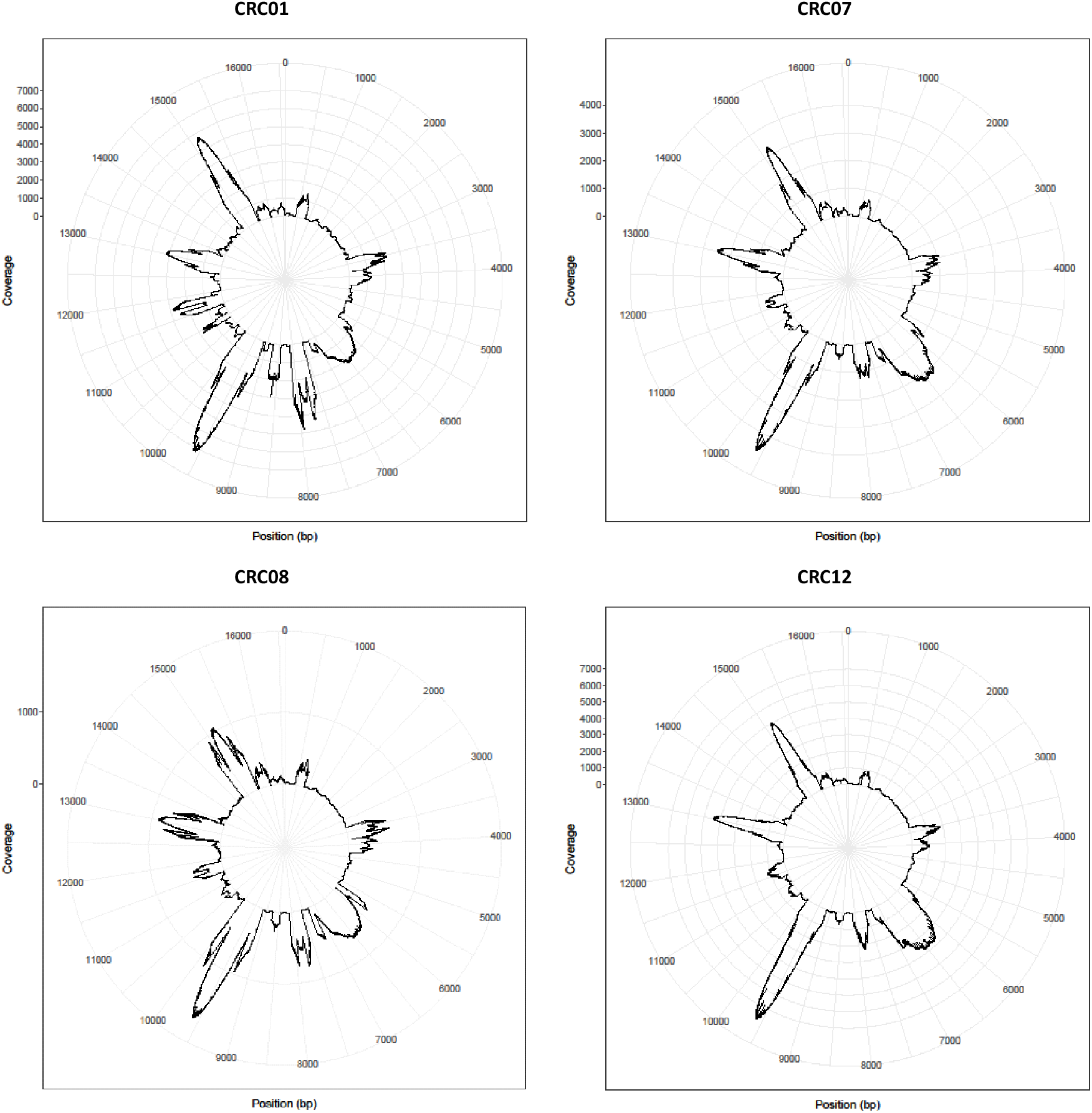
Coverage heterogeneity along the mitochondrial genome. The figure shows the average number of reads along the mtDNA sequence for each individual. Coverage for positions 1-4000 and 12001-16569 was calculated from the reads aligned to the shifted rCRS reference, while for positions 4001-12000, we calculated the coverage from the reads aligned to the rCRS reference.

**Figure S2.**
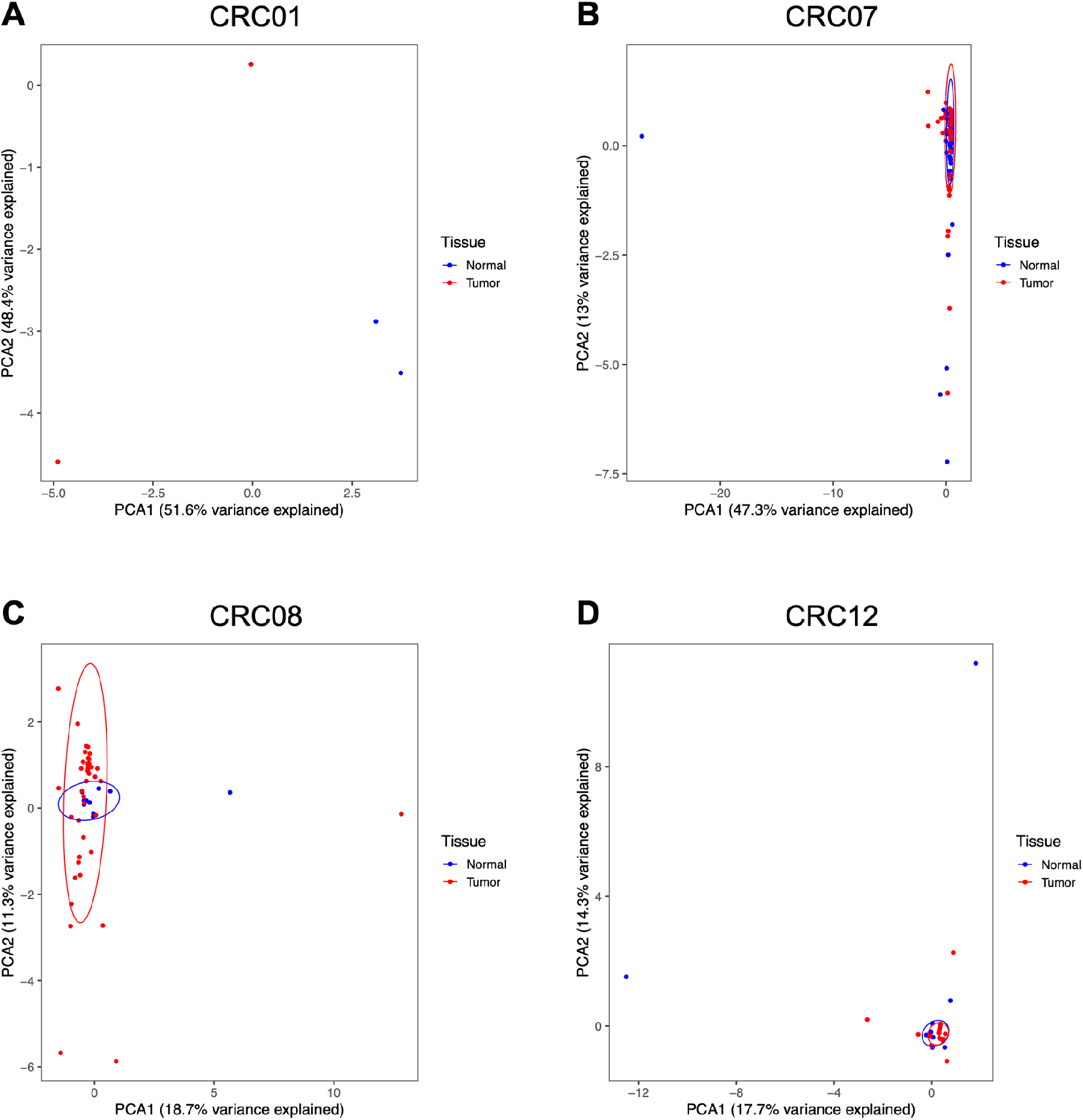
Principal component analysis (PCA) for normal and tumor cells. PCA for individuals CRC01 (A), CRC07 (B), CRC08 (C) and CRC12 (D). Colors correspond to normal (blue) and tumor (red) cells. Ellipses represent a Normal distribution with a probability of 95% for the respective color group.

**Figure S3.**
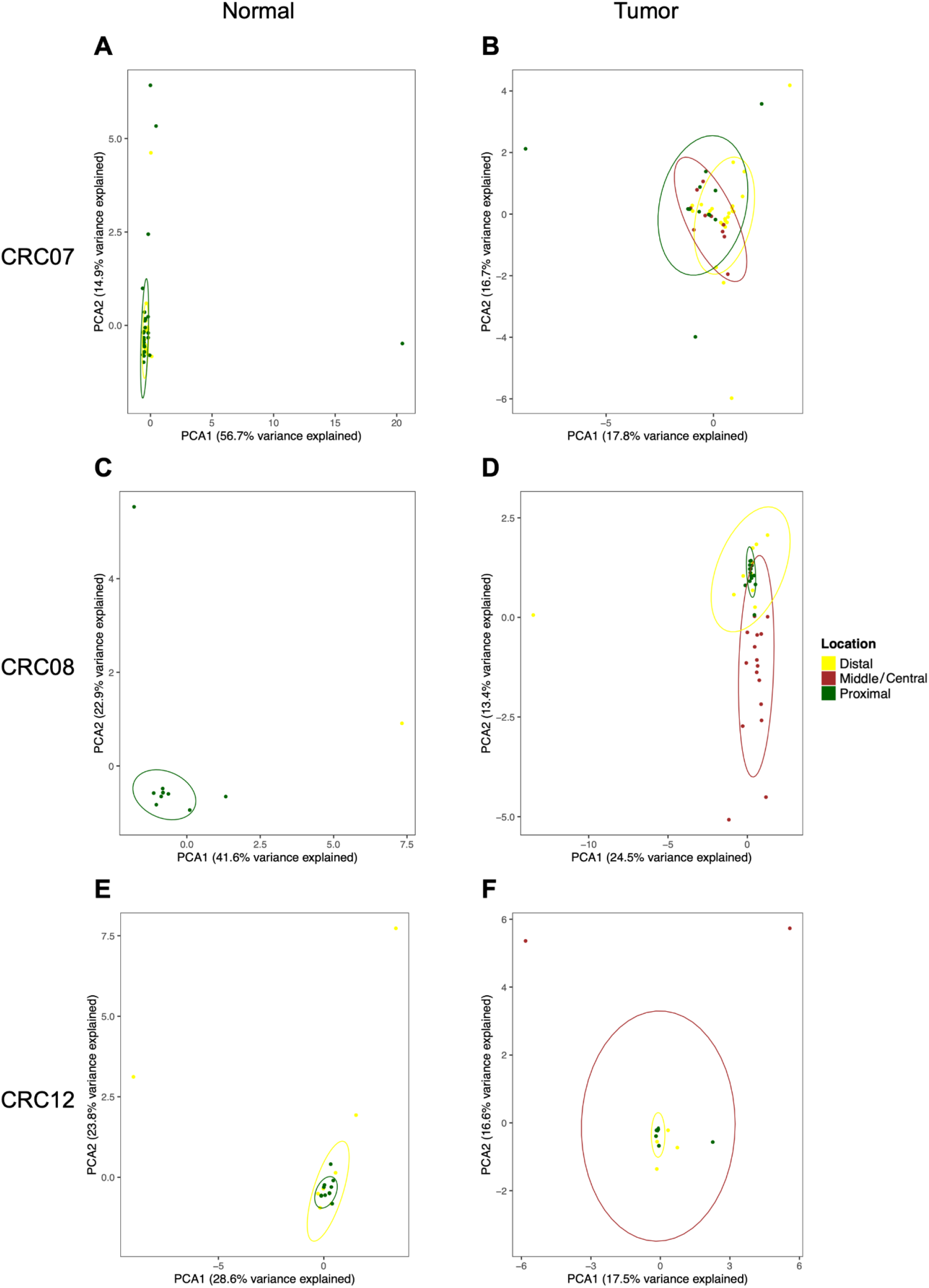
Principal component analysis (PCA) considering locations. PCA for individuals CRC07 (A and B), CRC08 (C and D), and CRC12 (E and F). Ellipses represent a Normal distribution with a probability of 95% for the respective color group. Left panels (A, C, and E) show only normal single-cells, and the right panels (B, D, and F) only tumor single-cells.

**Figure S4.**
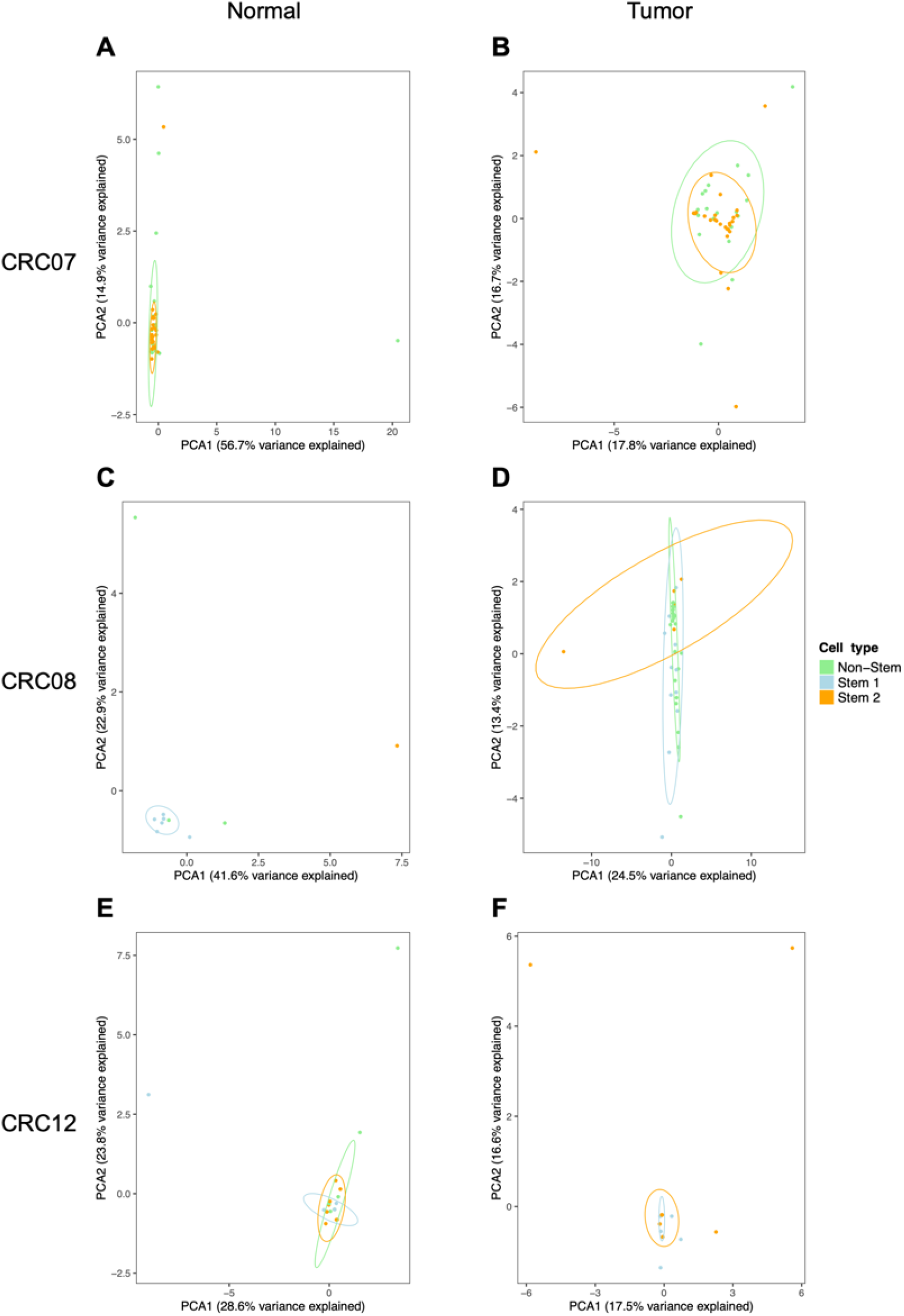
Principal component analysis (PCA) considering cell type. PCA for individuals CRC07 (A and B), CRC08 (C and D), and CRC12 (E and F). Colors according to cell type. Ellipses represent a Normal distribution with a probability of 95% for the respective color group. Left panels (A, C, and E) show only normal single-cells, and the right panels (B, D, and F) only tumor single-cells.

